# Understanding non-adherence to tuberculosis medications in India using urine drug metabolite testing: a cohort study

**DOI:** 10.1101/2021.01.12.21249665

**Authors:** Ramnath Subbaraman, Beena E. Thomas, J. Vignesh Kumar, Kannan Thiruvengadam, Amit Khandewale, S. Kokila, Maya Lubeck-Schricker, M. Ranjith Kumar, Gunjan Rahul Gaurkhede, Apurva Shashikant Walgude, J. Hephzibah Mercy, Jagannath Dattatraya Kumbhar, Misha Eliasziw, Kenneth H. Mayer, Jessica E. Haberer

## Abstract

**Background:** Suboptimal adherence to tuberculosis (TB) treatment is associated with increased disease recurrence and death. Little research has been conducted in India to understand TB medication non-adherence.

**Methods:** We enrolled adult drug-susceptible TB patients, about half of whom were people living with HIV (PLHIV), in Chennai, Vellore, and Mumbai. We conducted a single unannounced home visit to administer a survey assessing reasons for non-adherence and collect a urine sample that was tested for isoniazid content. We described patient-reported reasons for non-adherence and identified factors associated with non-adherence (negative urine test) using multivariable logistic regression. We also assessed the association between non-adherence and unfavorable treatment outcomes.

**Results:** Of 650 patients in the cohort, 77 (11.8%) had a negative urine test result. Non-adherence was independently associated with daily wage labor (aOR 3.1, CI: 1.3—7.7), smear-positive pulmonary disease (aOR 2.1, CI: 1.1—4.1), alcohol use (aOR 2.3, CI: 1.1—4.8), and spending 60 minutes or more picking up medication refills (aOR 9.1, CI: 1.8—45.4). PLHIV reported greater barriers to picking up medications than non-PLHIV. Among 167 patients who reported missing doses, common reasons reported included traveling away from home, forgetting, feeling depressed, and running out of pills. The odds of non-adherence was 3.8 (CI: 2.1—6.9) times higher among patients with unfavorable treatment outcomes compared to those with favorable outcomes.

**Conclusion:** Addressing structural and psychosocial barriers will be critical to improve TB treatment adherence in India. Urine isoniazid testing may help identify non-adherent patients to facilitate early intervention during treatment.

**Key points:** We evaluated adherence to tuberculosis medications in 650 Indian patients by conducting urine isoniazid tests collected during unannounced home visits. Structural barriers to collecting medication refills and psychosocial barriers emerged as the most important factors contributing to medication non-adherence.

## Introduction

Suboptimal adherence to active tuberculosis (TB) treatment is associated with increased disease recurrence and death [1–3]. A meta-analysis of recent clinical trials suggests that missing more than 10% of doses during treatment for drug-susceptible TB is associated with nearly six-fold increased odds of unfavorable outcomes, including disease recurrence [2]. Studies in programmatic conditions similarly reveal higher disease recurrence for TB patients with poor adherence [3]. A modeling study suggests that reducing missed doses could have larger epidemiological impact on reducing TB incidence than decreasing loss to follow-up during treatment in high burden countries [4].

Despite the importance of TB medication adherence, little research has evaluated factors contributing to non-adherence in high burden settings such as India, which has the world’s largest epidemic. The few previous Indian studies have identified clinical (e.g., medication adverse effects [5–7], symptom improvement [5]), psychosocial (e.g., alcohol use [5,6,8], stigma [5,6]), structural (e.g., distance from clinic [5–9], migration [5], work-related challenges [5,7,9]), and health system (e.g., non-cooperative staff [5], drug stockouts [8]) barriers contributing to non-adherence.

These previous studies have methodological limitations including small sample sizes [5–7]. Some studies reportedly assessing adherence actually measured loss to follow-up from treatment, rather than missed doses on a day-to-day basis, which is a behavior with potentially different contributing factors and clinical implications [10–13]. None of these studies employed rigorous approaches for measuring adherence, such as patient monitoring using digital adherence technologies (DATs) [14] or medication metabolite testing [15,16]—strategies that have been used to understand adherence in HIV and other conditions [17].

Moreover, prior adherence studies were conducted in the context of monitoring with directly observed therapy (DOT), usually using a facility-based approach, in which patients visited clinics where healthcare providers observed every dose. In recent years, concern has been growing regarding the patient and health system burden, ethical limitations, and effectiveness of DOT [14,18–21]. At the same time, in 2014, India’s National TB Elimination Program (NTEP) introduced daily medication dosing for all drug-susceptible TB patients, in place of the prior thrice-weekly dosing regimen [22]. This shift led to concerns that having patients visit clinics daily, rather than thrice weekly, might be too burdensome [22]. As a result, the NTEP has transitioned away from DOT and towards greater use of self-administered therapy (i.e., patients taking pills by themselves) or monitoring with 99DOTS, a cellphone-based DAT [22]. No recent studies have been conducted to understand causes of non-adherence to TB therapy in India with SAT or monitoring with DATs.

In this manuscript, we investigate non-adherence to TB medications using data from a cohort study conducted in three Indian cities among people living with HIV (PLHIV) and non-PLHIV. Adherence was assessed by testing patients’ urine for isoniazid content during unannounced home visits. Although the study’s primary goal, for which results have been reported [15], was to understand 99DOTS’ accuracy for measuring adherence, the collected data also provide rich insights into potential causes of non-adherence. The aims of the present study were to identify clinical, structural, and psychosocial factors associated with non-adherence; describe patient-reported reasons for non-adherence collected using a standardized questionnaire; and assess the association between non-adherence and TB treatment outcomes.

## Methods

### Ethics approvals

The study protocol was approved by ethics committees at the National Institute for Research in TB (NIRT, Chennai, India), Brigham and Women’s Hospital (Boston, USA), and Tufts University (Boston, USA).

### Study setting

To enroll a geographically diverse cohort, we recruited TB patients from three cities with a relatively high TB burden: Mumbai, Chennai, and Vellore [23,24], Nearly all patients in Mumbai were HIV-negative and recruited from 11 DOT centers with some of the highest patient volumes. All patients in Chennai and Vellore were HIV-positive and recruited from the five largest HIV antiretroviral therapy (ART) centers in these cities.

### Patient recruitment and data collection

From August 2017 to February 2019, we sequentially recruited adult patients (18 years of age and older) from the selected clinics who were taking treatment for drug-susceptible TB and being monitored with 99DOTS. We enrolled patients visiting clinics for treatment initiation and for collection of medication refills through the penultimate month of therapy. After exhausting the pool of patients who were already taking treatment, we continued to enroll patients who were starting treatment, but their unannounced home visit was randomly chosen to occur either in the intensive (first two months) or the continuation (last four months) phase.

Informed consent was obtained upon enrollment. Based on a socio-ecological framework [25], a baseline questionnaire was designed to collect data on demographics, socioeconomic status, structural barriers, and psychosocial challenges—including the brief alcohol use disorder identification test (AUDIT-C) [26]. Patients became eligible for a home visit three weeks after enrollment or after reaching the continuation phase, for those randomized to receive a home visit in that treatment phase. A random number generator was used to select the exact day of the home visit.

We attempted to minimize changes in adherence behavior in response to study interactions (“Hawthorne effect”). First, we conducted a single home visit with every patient under the assumption that they may be more likely to change their behavior with repeated visits. While this single visit limits understanding of adherence longitudinally for each patient, home visits for the cohort were distributed throughout the treatment course, providing reasonable representation of adherence for the population. Second, waiting at least three weeks for the home visit likely reduced the impact of short-term behavioral changes related to the initial study interaction. Third, the home visit was conducted without prior notice to minimize the likelihood of patients taking medications solely in anticipation of the visit.

During the home visit, we asked patients questions modified from the AIDS Clinical Trials Group adherence questionnaire to assess reasons for non-adherence [27]. Patients also provided a urine sample that was tested using IsoScreen, a validated assay in which reagents change color if isoniazid is present [28,29]. Approximately 100%, 83%, and 11% of patients will have positive Isoscreen results at 24, 48, and 72 hours, respectively, after last ingestion of isoniazid [28,29]. Positive test results comprise purple, blue, or green color changes, with purple or blue suggestive of medication ingestion within the last 24 hours and green suggestive of ingestion 24 to 48 hours before [28,29]. We defined “non-adherence” as consisting of a yellow test result (in comparison to green, blue, or purple results), indicating high likelihood of not having taken doses for 48 to 72 hours or longer. We defined “suboptimal adherence” as comprising yellow or green test results (in comparison to blue or purple results), indicating high likelihood of not having taken a dose for 24 hours or longer.

### Analyses

Using JMP Pro 15 (SAS Institute, Inc., Cary, NC), we performed univariable and multivariable logistic regression analyses to identify clinical, structural, and psychosocial factors associated with non-adherence, as well as with suboptimal adherence. All demographic and clinical factors were retained in the multivariable model by design. Other factors were retained if significant at p<0.2 in the univariable analyses. Modifications made to the model to remediate multicollinearity are described in the supplementary appendix. We included alcohol use as a binary variable comparing those with moderate or high use (AUDIT-C score of 1 or greater) to no alcohol use (score of 0). For additional insights into non-adherence, we described the proportion of patients reporting various reasons for non-adherence in the home visit questionnaire.

Finally, we evaluated whether the urine test result was associated with TB treatment outcomes. Using logistic regression, we assessed the association between non-adherence and death, loss to follow-up, and treatment failure as separate outcomes, and with these outcomes aggregated as “unfavorable outcomes.” We obtained outcomes from patients’ treatment cards through February 2019, when the study ended, and later from Nikshay (the NTEP’s electronic medical record) in September 2020. We analyzed the association between non-adherence and treatment outcomes separately using each data source. Details on each data source are in the supplementary appendix.

## Results

### Descriptive statistics

Of 832 patients who met eligibility criteria, 84 (10%) did not consent for the study or could not enroll because family members collected their medications. Of 748 patients who enrolled, we could not complete home visits for 98 (13%), despite three attempts (supplementary appendix, Figure S1).

Of 650 patients in the final analysis, 77 (11.8%) were non-adherent and 116 (17.8%) had suboptimal adherence. More than three-quarters had a household monthly income of <15,000 Indian rupees (INR; approximately <200 US dollars [USD]), and more than two-thirds had not completed high school (Table 1). Most patients were new (formerly “category 1”), had smear-positive pulmonary TB, and were in the continuation phase at the time of the home visit. Few were able to walk or bicycle to clinic; most spent 60 minutes or more collecting medication refills.

**Table 1.** Descriptive statistics for the cohort and factors associated with non-adherence to TB medications (N=650)

| Covariates | Descriptive Statistics |  | Univariable findings |  | Multivariable findings |  |
| --- | --- | --- | --- | --- | --- | --- |
|  | Proportion of overall sample in given category<sup>a</sup> | Proportion with non-adherence<sup>b</sup> | Odds ratio (95% confidence interval) | p-value | Odds ratio (95% confidence interval) | p-value |
|  | N (%) | n (%) |  |  |  |  |
| **DEMOGRAPHIC FACTORS** |  |  |  |  |  |  |
| **Gender** |  |  |  |  |  |  |
| Female | 271 (41.7) | 28 (10.3) | Ref |  | Ref |  |
| Male | 379 (58.3) | 49 (12.9) | 1.3 (0.8—2.1) | 0.31 | 0.9 (0.4—1.8) | 0.74 |
| **Age** |  |  |  |  |  |  |
| 18-24 | 156 (24.0) | 9 (5.8) | Ref |  | Ref |  |
| 25-34 | 145 (22.3) | 30 (20.7) | 4.3 (1.9—9.3) | 0.0003* | 2.4 (0.99—6.0) | 0.052 |
| 35-44 | 176 (27.1) | 21 (11.9) | 2.2 (0.98—5.0) | 0.056 | 0.6 (0.2—1.7) | 0.35 |
| ≥45 | 173 (26.2) | 17 (9.8) | 1.8 (0.8—4.1) | 0.178 | 0.5 (0.2—1.4) | 0.17 |
| **Income quartiles** |  |  |  |  |  |  |
| <5,000 | 98 (15.1) | 16 (16.3) | Ref |  | Ref |  |
| 5000—9,999 | 223 (34.3) | 27 (12.1) | 0.7 (0.4—1.4) | 0.30 | 0.7 (0.3—1.5) | 0.33 |
| 10,000—14,999 | 186 (28.6) | 23 (12.4) | 0.7 (0.4—1.4) | 0.36 | 1.2 (0.5—2.9) | 0.61 |
| ≥15,000 | 143 (22.0) | 11 (7.7) | 0.4 (0.2—0.97) | 0.04* | 0.6 (0.2—1.7) | 0.39 |
| **Educational level** |  |  |  |  |  |  |
| No formal education | 116 (17.9) | 17 (14.7) | Ref |  | Ref |  |
| 6 or fewer years | 148 (22.8) | 23 (15.5) | 1.1 (0.5—2.1) | 0.84 | 1.3 (0.6—2.7) | 0.57 |
| 7 to 12 years | 180 (27.7) | 19 (10.6) | 0.7 (0.3—1.4) | 0.29 | 0.9 (0.4—1.9) | 0.70 |
| Higher secondary completed or college | 206 (31.7) | 18 (8.7) | 0.6 (0.3—1.1) | 0.10 | 0.8 (0.3—1.9) | 0.60 |
| **Occupation** |  |  |  |  |  |  |
| Self employed | 137 (21.1) | 12 (8.8) | Ref |  | Ref |  |
| Employed in government or private sector | 130 (20.0) | 16 (12.3) | 1.5 (0.7—3.2) | 0.35 | 2.1 (0.9—4.9) | 0.10 |
| Laborer on daily wages | 84 (12.9) | 16 (19.0) | 2.5 (1.1—5.5) | 0.03* | 3.1 (1.3—7.7) | 0.01* |
| Unemployed | 133 (20.5) | 23 (17.3) | 2.2 (1.04—4.6) | 0.04* | 2.0 (0.8—4.7) | 0.12 |
| Housewife or student | 166 (25.5) | 10 (6.0) | 0.7 (0.3—1.6) | 0.36 | 1.0 (0.3—3.1) | 0.96 |
| **Marital Status** |  |  |  |  |  |  |
| Never married | 169 (26.0) | 17 (10.1) | Ref |  | Ref |  |
| Married or living with partner | 411 (63.2) | 48 (11.7) | 1.2 (0.7—2.1) | 0.57 | 1.4 (0.7—2.8) | 0.32 |
| Divorced, separated, or widowed | 70 (10.8) | 12 (17.1) | 1.8 (0.8—4.1) | 0.13 | 1.6 (0.6—4.5) | 0.34 |
| **CLINICAL FACTORS** |  |  |  |  |  |  |
| **Phase of Therapy** |  |  |  |  |  |  |
| Intensive phase | 217 (33.4) | 25 (11.5) | Ref |  | Ref |  |
| Continuation phase | 433 (66.6) | 52 (12.0) | 1.0 (0.6—1.7) | 0.86 | 1.5 (0.8—2.6) | 0.17 |
| **Category of TB** |  |  |  |  |  |  |
| New | 504 (77.5) | 55 (10.9) | Ref |  | Ref |  |
| Previously treated | 146 (22.5) | 22 (15.1) | 1.4 (0.9—2.5) | 0.17 | 1.6 (0.9—2.9) | 0.12 |
| **Type of TB** |  |  |  |  |  |  |
| Extrapulmonary | 202 (31.1) | 15 (7.4) | Ref |  | Ref |  |
| Smear-negative pulmonary | 77 (11.9) | 11 (14.3) | 2.1 (0.9—4.8) | 0.08 | 2.2 (0.9—5.5) | 0.097 |
| Smear-positive pulmonary | 371 (57.1) | 51 (13.8) | 2.0 (1.1—3.6) | 0.03* | 2.1 (1.1—4.1) | 0.03* |
| **HIV co-infection** |  |  |  |  |  |  |
| No | 347 (53.4) | 26 (7.5) | Ref |  | Ref |  |
| Yes | 303 (46.6) | 51 (16.8) | 2.5 (1.5—4.1) | 0.0003* | 1.6 (0.6—4.1) | 0.33 |
| **STRUCTURAL FACTORS** |  |  |  |  |  |  |
| **Transport mode to clinic** |  |  |  |  |  |  |
| Walking or bicycle | 234 (36.0) | 13 (5.6) | Ref |  |  |  |
| Motorcycle or car | 44 (6.8) | 5 (11.4) | 2.2 (0.7—6.5) | 0.15 |  |  |
| Authorickshaw or taxi | 131 (20.2) | 14 (10.7) | 2.0 (0.9—4.5) | 0.08 |  |  |
| Public transportation | 241 (37.1) | 45 (18.7) | 3.9 (2.0—7.4) | <0.0001* |  |  |
| **Money spent to collect medications** |  |  |  |  |  |  |
| INR 0—24 | 233 (35.9) | 17 (7.3) | Ref |  |  |  |
| INR 25—49 | 111 (17.1) | 10 (9.0) | 1.3 (0.6—2.8) | 0.58 |  |  |
| INR 50—75 | 111 (17.1) | 17 (15.3) | 2.3 (1.1—4.7) | 0.02* |  |  |
| INR >75 | 195 (30.0) | 33 (16.9) | 2.6 (1.4—4.8) | 0.003* |  |  |
| **Time spent to collect medications** |  |  |  |  |  |  |
| <30 minutes | 110 (16.9) | 2 (1.8) | Ref |  | Ref |  |
| 30 to 59 minutes | 191 (29.4) | 19 (9.9) | 6.0 (1.4—26.1) | 0.02* | 6.2 (1.4—28.1) | 0.018* |
| ≥60 minutes | 349 (53.7) | 56 (16.0) | 10.3 (2.5—43.0) | 0.001* | 9.1 (1.8—45.4) | 0.007* |
| **PSYCHOSOCIAL FACTORS** |  |  |  |  |  |  |
| **Current tobacco use** |  |  |  |  |  |  |
| No | 540 (83.0) | 59 (10.9) | Ref |  |  |  |
| Smokeless tobacco only | 51 (7.9) | 4 (7.8) | 0.7 (0.2—2.0) | 0.50 |  |  |
| Cigarette or beedi use | 59 (9.1) | 14 (23.7) | 2.5 (1.3—4.9) | 0.006* |  |  |
| **Probable alcohol use** |  |  |  |  |  |  |
| No alcohol use | 591 (90.9) | 61 (10.3) | Ref |  | Ref |  |
| Any alcohol use | 59 (9.1) | 16 (27.1) | 3.2 (1.7—6.1) | 0.0003* | 2.3 (1.1—4.8) | 0.03* |

INR=Indian rupees.
<sup>a</sup>Represents the number of participants in a category divided by the overall cohort sample of 650—e.g., there are 271/650 females in the cohort.
<sup>b</sup>Represents the number of participants with non-adherence in a given category—e.g., 28/271 females were non-adherent.

### Factors associated with non-adherence and sub-optimal adherence

In the multivariable model with non-adherence as the outcome, individuals who were laborers on daily wages, were smear-positive pulmonary TB patients, spent 30 or more minutes collecting medications, or used alcohol were at significantly increased odds of being non-adherent (Table 1). In the multivariable model with suboptimal adherence as the outcome, patients who were 35 or older were at decreased odds of being sub-optimally adherent. Patients who were daily wage laborers, spent 30 or more minutes collecting medications, or used alcohol were at significantly increased odds of being sub-optimally adherent (supplementary appendix, Table S2).

### Differences in structural challenges facing PLHIV and non-PLHIV

Compared to non-PLHIV, PLHIV were more likely to take public transportation, spend ≥240 minutes, and spend more than INR 75 (USD 1) per clinic visit to collect medication refills (Table 2).

**Table 2.**
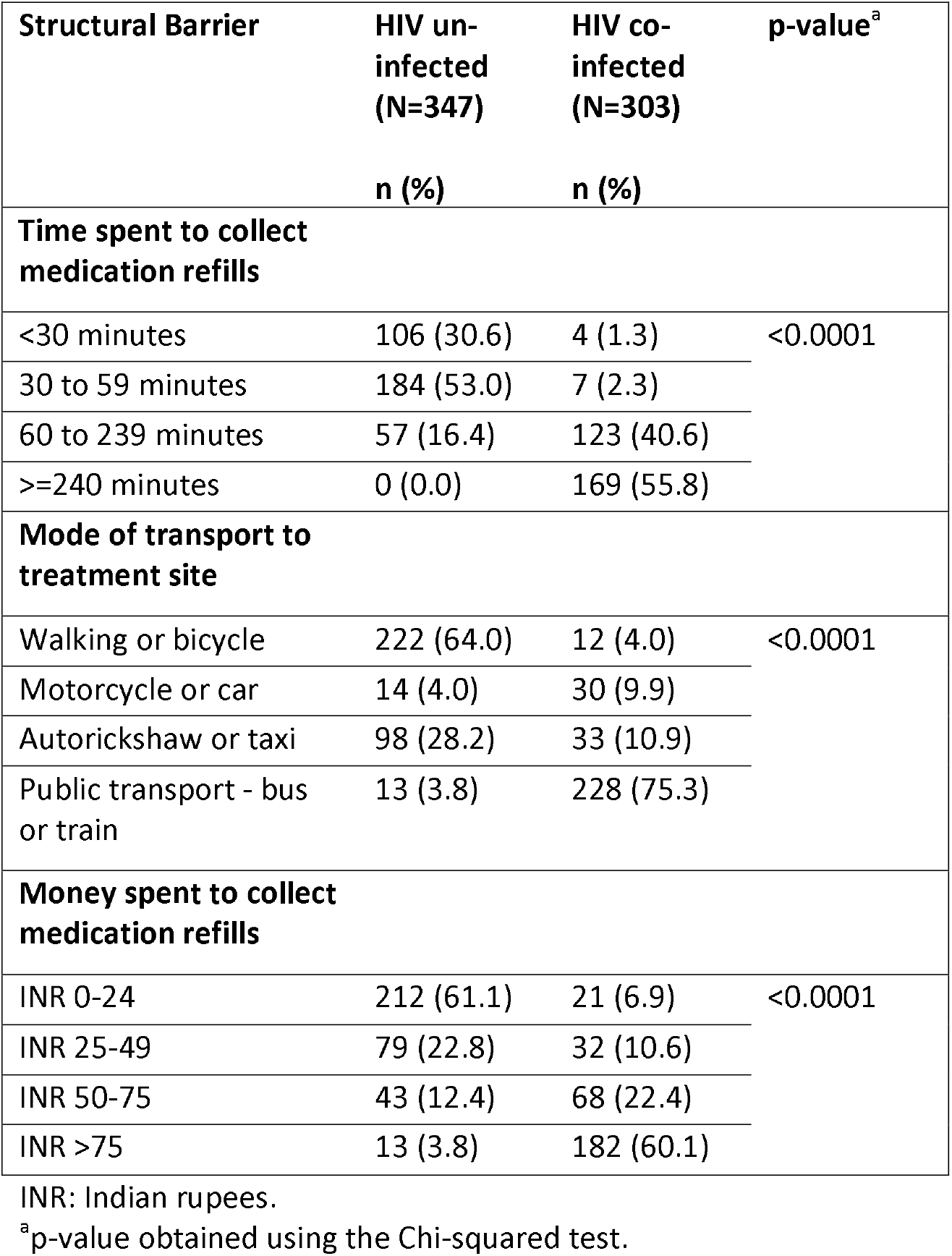
Association of being a person living with HIV with structural barriers in getting to the clinic to pick up TB medication refills.

### Patient-reported reasons for non-adherence

Of 650 patients who answered the home visit survey, 167 (25.7%) reported missing at least one medication dose during therapy. Of these, more than one-fifth reported traveling or being away from home, forgetting to take medications, feeling depressed, or running out of pills as contributing to non-adherence (Table 3). See supplementary appendix for details on why patients ran out of pills and medication adverse effects that negatively impacted adherence.

**Table 3:** Patient-reported reasons for missing tuberculosis (TB) medication doses (N=167)

| Reason for missing doses<sup>a</sup> | Proportion of patients reporting this problem<sup>b</sup> | Frequency with which patients were affected by this problem |  |  |
| --- | --- | --- | --- | --- |
|  |  | Rarely n(%) | Sometimes n(%) | Often n(%) |
| Traveling or being away from home | 67 (40.1%) | 37 (22.2%) | 19 (11.4%) | 11 (6.6%) |
| Simply forgot | 50 (29.9%) | 28 (16.8%) | 11 (6.6%) | 11 (6.6%) |
| Felt depressed | 39 (23.3%) | 16 (9.6%) | 16 (9.6%) | 7 (4.2%) |
| Ran out of pills | 35 (21.0%) | 29 (17.4%) | 3 (1.8%) | 3 (1.8%) |
| Wanted to avoid medication adverse effects | 29 (17.4%) | 15 (9.0%) | 8 (4.8%) | 6 (3.6%) |
| Reduced motivation because TB symptoms improved | 19 (11.4%) | 7 (4.2%) | 5 (3.0%) | 7 (4.2%) |
| Had too many pills to take for different conditions | 17 (10.2%) | 9 (5.4%) | 4 (2.4%) | 4 (2.4%) |
| Did not want others to notice me taking medication | 16 (9.6%) | 12 (7.2%) | 2 (1.2%) | 2 (1.2%) |

<sup>a</sup>Patients could report more than one reason for missing doses.
<sup>b</sup>Represents the proportion of individuals reporting a given problem over the total number in the cohort who reported having ever missed medication doses; for example, 67/167 study participants reported traveling or being away from home as a reason for missing doses.

### Association between non-adherence to medications and TB treatment outcomes

Using Nikshay data, non-adherence was significantly associated with subsequent loss to follow-up and with the composite outcome of treatment failure, regimen change, and transfer of care out of district (Table 4). Patients with any unfavorable outcome were at 3.8 (95%CI 2.1—6.9) increased odds of being non-adherent compared to those with favorable outcomes. The same analysis conducted using outcomes from treatment cards yielded similar findings (supplementary appendix, Table S3).

**Table 4.**
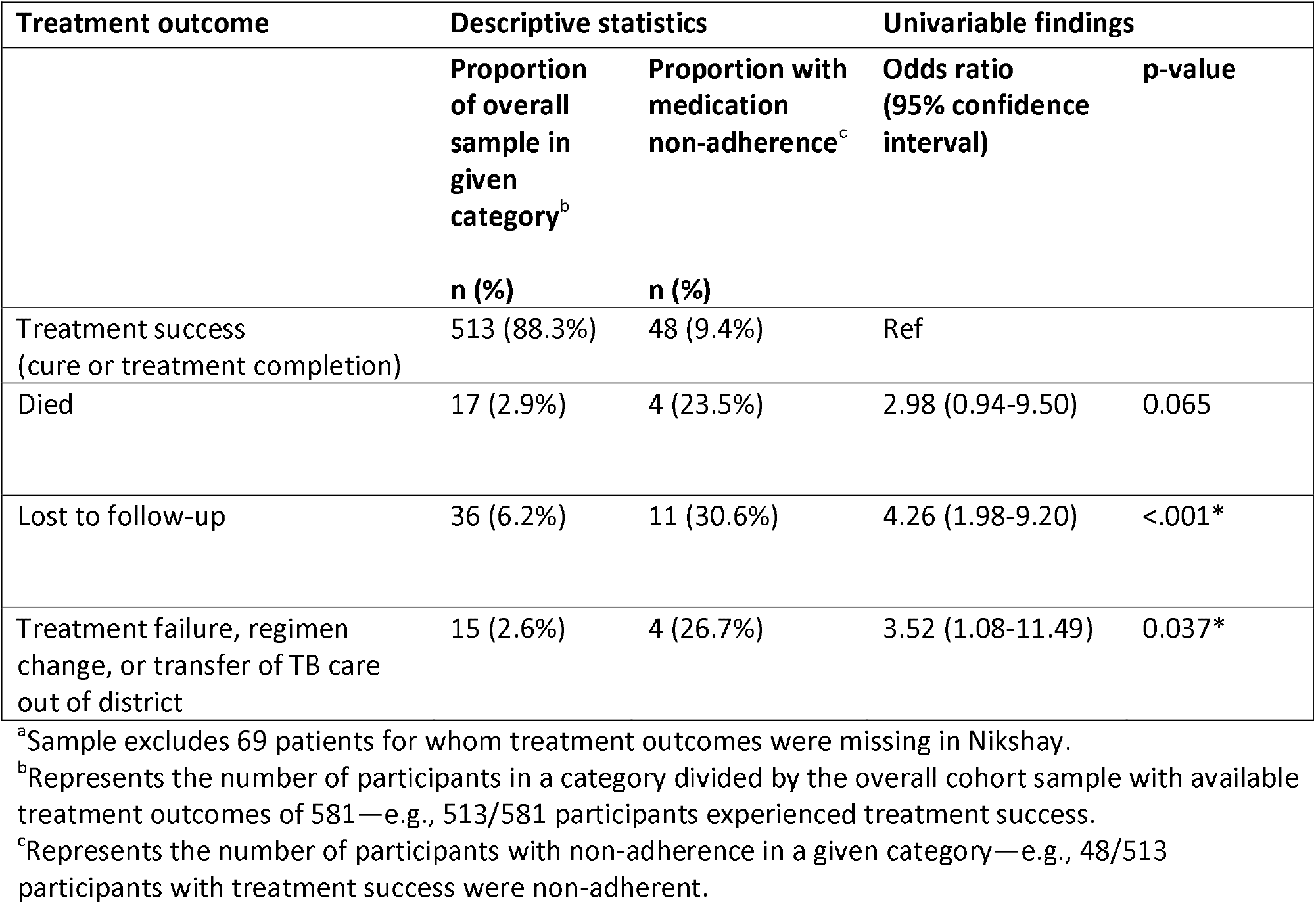
Association between non-adherence to tuberculosis (TB) medications and treatment outcomes reported in India’s National TB Elimination Program Nikshay system for the cohort (N=581)^a^.

## Discussion

In this cohort study, we used survey questions and urine metabolite testing, a rigorous marker of isoniazid ingestion, to identify factors associated with non-adherence to drug-susceptible TB treatment in India. We found that medication non-adherence is a complex problem, involving structural, psychosocial, and clinical barriers that may be amenable to intervention.

Structural barriers may be a central challenge leading to non-adherence. The adjusted odds of non-adherence increased considerably for patients who spent 30 minutes or more collecting medication refills—a factor that was collinear with other structural barriers (i.e., transportation mode and money spent collecting medications). These structural barriers triangulate with the finding in the home visit survey that more than one-fifth of patients reported running out of pills as a reason for missing doses. Similar to findings in the HIV literature, many patients attributed shortage of pills to transportation difficulties or other challenges collecting medications [30].

PLHIV disproportionately experienced these structural barriers compared to non-PLHIV, likely due to recent changes in TB care delivery, in which PLHIV now collect TB and HIV medication refills concurrently, or in a “single window,” at ART centers. Previously, PLHIV collected TB medications at DOT centers located near their homes, similar to non-PLHIV. While DOT centers are decentralized in primary health centers, there are fewer ART centers in India that are often located in tertiary hospitals. Although “one-stop shopping” seems to be a laudable goal, creation of new barriers through longer commutes to ART centers may attenuate the benefits of consolidated pharmacy services. Our findings may explain results of a study in Karnataka, India, in which treatment outcomes were worse for PLHIV receiving TB treatment at ART centers that had implemented the single window system, as compared to PLHIV receiving treatment at DOT centers [31].

Interventions to address structural barriers may include providing travel vouchers to help patients get to clinic or delivering medication refills to patients. For example, recent studies have evaluated the benefits of HIV adherence clubs, in which PLHIV meet locally to discuss their care and collect ART, making it easier for PLHIV to get their medications while providing them with counseling [32]. Such interventions merit evaluation in TB patients in India.

Laborers receiving daily wages had higher non-adherence. This finding remained significant after adjusting for household income, suggesting that this increased risk may relate to job constraints, such as loss of wages when they go to collect medications, rather than income poverty more generally. Home delivery of medications may benefit patients working in such jobs. Many patients reported traveling or being away from home as a reason for missing doses, which may also reflect job-related structural constraints, a barrier also reported in the HIV literature [33]. Future research should examine how adherence and medication access can be supported during travel.

Psychosocial factors also emerged as important barriers to adherence. Alcohol use was significantly associated with non-adherence, consistent with findings from previous Indian studies showing poorer TB treatment outcomes in patients with alcohol use disorder [34,35]. While we did not formally measure depression, more than one-fifth of patients who reported missing doses described feeling depressed as a major reason. A recent systematic review found that depression is associated with increased loss to follow-up and death—but not medication non-adherence—during TB treatment; however, the included studies had suboptimal measures of adherence [36]. Future studies using rigorous measures of both depression and adherence, such as the urine testing we used here, may help to understand whether non-adherence mediates the association between depression and treatment outcomes. A few patients reported missing doses because they did not want others to notice them taking medications, highlighting stigma as another barrier to adherence [37,38].

Clinical factors may also contribute to non-adherence. Smear-positive pulmonary TB patients had higher adjusted odds of non-adherence. These patients have poorer treatment outcomes on average compared to smear-negative and extrapulmonary TB patients in the NTEP [39]. Our finding raises a question of whether poorer treatment outcome in smear-positive patients may be mediated not only by their greater disease severity but also by higher non-adherence. Desire to avoid medication adverse effects was also reported as a reason for missing doses, consistent with findings from previous Indian studies [40]. Healthcare providers often believe that improvement in patients’ TB symptoms later in therapy is a major cause of non-adherence; however, only a small proportion of patients described symptom improvement as a reason for missing doses. Enhanced counseling may help address patient concerns about adverse effects while also increasing motivation to adhere to therapy even after symptoms improve.

Many patients said they simply “forgot” to take medications. While often thought of as a cognitive problem, forgetfulness is also shaped by structural and psychosocial barriers. Depression and alcohol use can reduce cognitive function, increasing the chance patients will forget to take pills, while structural barriers to collecting medications may make it more likely that patients will prioritize other tasks. One rationale for using DATs is that reminders provided by these technologies might reduce forgetfulness; however, these reminders do not address these contextual barriers that may contribute to patients forgetting medication doses.

Despite only conducting a single urine isoniazid test for each patient, a negative test result was strongly associated with unfavorable treatment outcomes, especially loss to follow-up. This finding highlights possible benefits of urine testing not only for research, but also for identifying patients at risk for poor outcomes in routine care. Conducting multiple urine tests throughout therapy might facilitate more robust prediction of long-term patient outcomes. However, further research is needed to understand whether urine testing might be acceptable to patients as part of routine care, evaluate whether tests conducted during clinic visits are as accurate as those collected during unannounced home visits [17], and ensure that testing does not have the unintended consequence of leading healthcare providers to stigmatize non-adherent patients.

One study limitation is that our cohort may not be representative of the overall TB patient population at our clinical sites, because some patients did not consent for the home visit and others could not consent as family members collected their medications. Adherence may be higher in our study population, because enrolled patients may have been more motivated to adhere than patients who declined to enroll. We had a modest number of outcomes (i.e., patients with negative urine tests), which may have limited statistical power to identify factors associated with non-adherence. Future studies involving a larger patient sample, with multiple urine tests on each patient, might identify additional factors associated with non-adherence.

## Conclusion

In this study of patients with drug-susceptible TB in India, we found non-adherence to be a complex problem shaped by a variety of structural, psychosocial, and clinical barriers. Of these challenges, structural barriers—in particular, transportation challenges and spending considerable time and money to collect medications—had the strongest association with non-adherence, particularly for PLHIV. Psychosocial barriers such as alcohol use, depression, and TB-associated stigma also emerged as major problems contributing to non-adherence.

Our findings suggest a need to facilitate easier access to medication refills and to develop counseling interventions that can simultaneously address depression, substance use disorders, and stigma. We also highlight drug metabolite testing as a useful approach for measuring adherence for TB patients taking self-administered or DAT-monitored therapy. Future research should assess whether urine testing can be used to improve adherence in routine clinical care and on developing multi-component interventions to address the diverse challenges contributing to non-adherence to TB medications in high burden countries.

## Data Availability

Requests for the de-identified dataset can be made by contacting Dr. Beena Thomas, although access to these data may be limited by NIRT/Indian Council of Medical Research policies.

## Acknowledgements

We thank all the participants who took part in this study. We acknowledge the immense contribution of all the field investigators for their efforts in collecting data. The Mumbai team included Yogesh Nivrutti Salve, Kamble Rakesh Suresh, Shweta Shantaram Bagade, and Meena Atul Kamble. The Tamil Nadu team included C. Jeganathan, S. Yokeshwaran, K. Sathiyamoorthy, E. Michael Raj, P. Brindadevi, K. Jegan, B.Sathyan Raj Kumar, and Mariyamma Paul.

