## Supplementary Appendix for "Understanding non-adherence to tuberculosis medications in India using urine drug metabolite testing: a cohort study"

### Methods

*Patient recruitment and data collection*

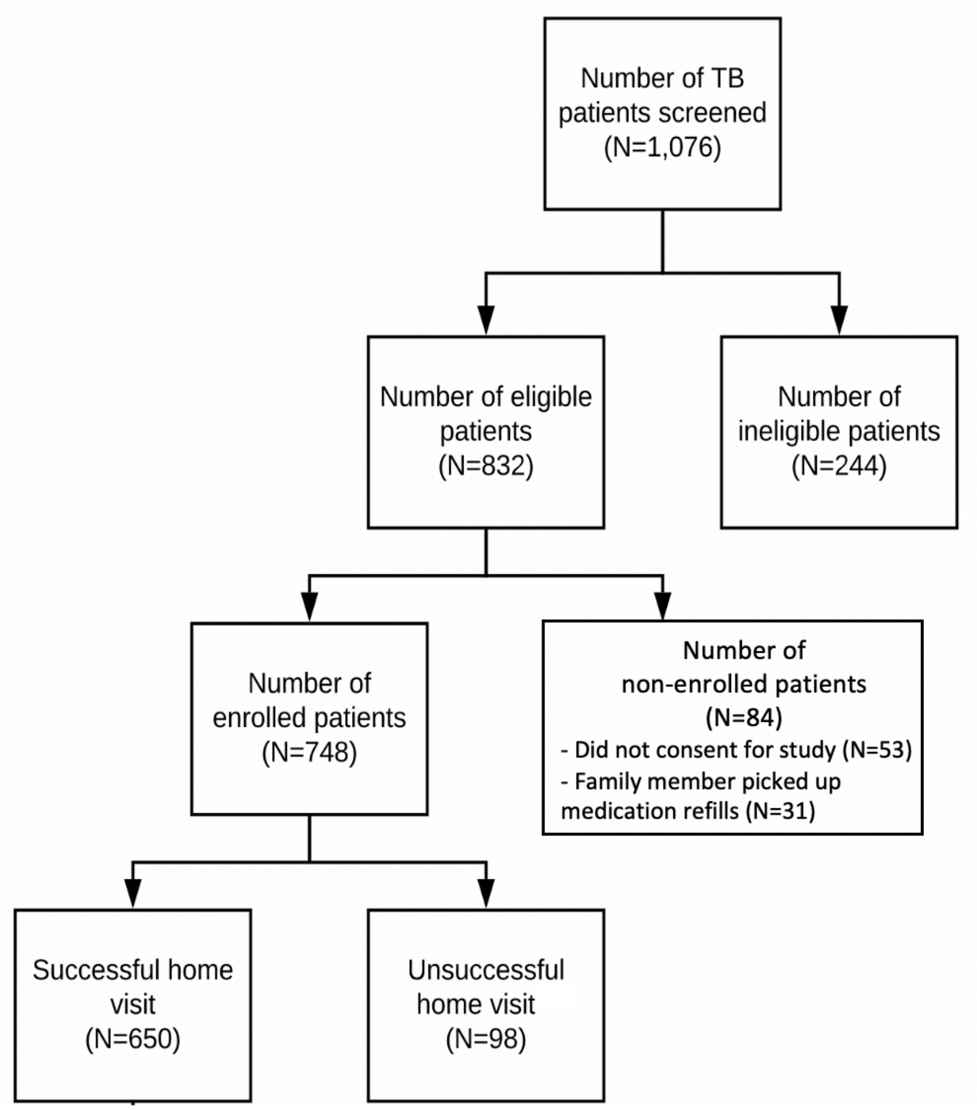


**Figure S1: Patients screened, enrolled, and home visits completed in the study cohort.** Patients with unsuccessful home visits were only classified as such after three unsuccessful home visit attempts by the research team.

*Addressing multicollinearity in the regression analyses*

When conducting regression diagnostics, HIV status was observed to have a high degree of multicollinearity (variance inflation factor >5) with mode of transport, money spent collecting medication refills, and time spent collecting medication refills. By removing mode of transport and money spent collecting medication refills from the model, we were able to reduce multicollinearity and retain HIV status and time spent collecting medication refills, which is the structural barrier with the strongest association with non-adherence. Similarly, given high multicollinearity between alcohol and tobacco use, we removed tobacco use from our model, because alcohol use seemed more likely to contribute to behavioral changes (e.g., inebriation, inattention, etc.) that could contribute to non-adherence.

*Analysis of the association between non-adherence and treatment outcomes – description of the two data sources*

Treatment card outcomes were collected directly from the paper-based treatment cards maintained by TB staff at DOT centers. These are standardized cards that include basic demographic information about the patients, information on medication refills collected, and treatment outcomes. Possible treatment outcomes recorded on treatment cards are cure, treatment completion, loss to follow-up (formerly default), death, treatment regimen change, transferred out, and still on treatment (Table S1).

Patient outcomes from treatment cards are then entered into Nikshay, the NTEP's electronic medical record, at a later date, usually by personnel at a centralized location such as a district or city TB office. Because data from paper treatment cards are sometimes entered into Nikshay weeks or months later, we waited to obtain data from Nikshay until September 2020. Possible treatment outcomes recorded in the Nikshay database are cure, treatment completion, loss to follow up, death, treatment regimen change, treatment failure, null, not evaluated, and closed due to inactivity (Table S1). We interpreted the last three of these outcomes (i.e., null, not evaluated, and closed due to inactivity) as representing missing data on treatment outcomes in Nikshay.

**Table S1. Comparison of treatment card and Nikshay data sources**

| | **Treatment card outcomes** | **Nikshay outcomes** |
|---|---|---|
| **Description of data source** | Standardized paper-based cards maintained by TB staff at DOT centers that include basic demographic information about the patients, information on medication refills collected, and treatment outcomes. | Patient outcomes from treatment cards entered into Nikshay, the NTEP's electronic medical record, at a later date, usually by personnel at a centralized location such as a district or city TB office. |
| **Possible treatment outcomes** | • Cure<br>• Treatment completion<br>• Loss to follow-up<br>• Death<br>• Treatment regimen change<br>• Transferred out<br>• On treatment | • Cure<br>• Treatment completion<br>• Loss to follow-up<br>• Death<br>• Treatment regimen change<br>• Treatment failure<br>• Null<br>• Not evaluated<br>• Closed due to inactivity |
| **Limitations of data source** | Research team was not able to collect final outcomes from treatment cards for patients who were still on treatment when the study ended in February 2019. | Treatment outcomes were missing in Nikshay for 133 (21%) patients in our cohort. If available, we substituted outcomes collected from paper treatment cards for the missing data. Four (0.6%) patients had treatment outcomes in Nikshay that were discordant with what was reported on patients' treatment cards. In these cases, Nikshay outcomes were substituted with treatment card outcomes. |
| **Approach to handling missing outcomes** | For the 269 (41%) patients who were still on treatment, the category of "on treatment" was included as a treatment outcome in the regression analysis. | For 69 (11%) patients with remaining missing treatment outcomes, these patients were excluded from the patient sample and therefore were not included in the regression analysis. |

Our research team collected treatment outcomes directly from the paper treatment cards through the end of February 2019, when our field research ended. Because our research team was able to collect this information directly from the paper-based records on which patient information is initially recorded, we believe that the treatment card-based outcomes are highly accurate. However, a limitation of the treatment card data is that we were not able to collect final outcomes from treatment cards for 269 (41%) patients, because they were still on treatment when the study ended (Table S1).

Due to this limitation of the treatment card data, we also conducted analyses using data from Nikshay, which recorded patient treatment outcomes beyond the end of our field research. Unfortunately, as of September 2020, treatment outcomes were not entered in Nikshay for 133 (20.5%) patients in our cohort. For these patients, we substituted outcomes collected from paper treatment cards, if available. Notably, 69 (10.6%) patients still had missing outcomes and were excluded from our analysis using Nikshay data. In rare cases when outcomes reported on the treatment cards conflicted with the Nikshay data, we substituted the outcome recorded by Nikshay with the treatment card outcome, because we believe the treatment card data were less likely to be incorrect due to data entry errors. Only 4 (0.6%) patients had treatment outcomes in Nikshay that were discordant with what was reported on patients' treatment cards (Table S1).

### Results

*Factors associated with non-adherence to TB medications – univariable analysis findings*

In univariable analyses with non-adherence as the outcome, individuals who were 25-34 years old, laborers on daily wages, unemployed, smear-positive pulmonary TB patients, HIV co-infected, taking public transportation to collect medications, spending 50 or more rupees to collect medications, taking 30 or more minutes to collect medications, using cigarettes or beedis, and using alcohol were at significantly increased odds of being non-adherent. Patients with a household monthly income 15,000 or more rupees were at significantly decreased odds of being non-adherent (main manuscript, Table 1).

*Factors associated with suboptimal adherence to TB medications – univariable analysis findings and table with findings from univariable and multivariable analyses*

In univariable analyses with suboptimal medication adherence as the outcome, patients who are 25-34 years old, laborers on daily wages, housewives or students, divorced/separated/widowed, take public transportation to collect their medications, spend 50 or more rupees to collect their medication, take 30 or more minutes to collect their medication, and use alcohol are statistically significantly more likely to be sub-optimally adherent. Patients with an income quartile greater than 15,000 rupees and who completed 7-12 years of education are significantly less likely to be sub-optimally adherent (Table S2). Findings of the multivariable analysis are shown in Table S2 and described in the main text of the manuscript.

Table S2. Factors associated with suboptimal adherence to TB medications (N=650)

| Covariates | Descriptive Statistics | Univariable findings | | Multivariable findings | |
|---|---|---|---|---|---|
| | Proportion of suboptimal adherence[a]<br><br>n (%) | Odds ratio (confidence interval) | p-value | Odds ratio (confidence interval) | p-value |
| **DEMOGRAPHIC FACTORS** | | | | | |
| **Gender** | | | | | |
| Female | 49 (18.1) | Ref | | Ref | |
| Male | 67 (17.7) | 1.0 (0.6—1.5) | 0.90 | 0.9 (0.5—1.6) | 0.71 |
| **Age** | | | | | |
| 18-24 | 20 (12.8) | Ref | | Ref | |
| 25-34 | 36 (24.8) | 2.2 (1.2—4.1) | 0.01* | 1.2 (0.6—2.5) | 0.56 |
| 35-44 | 30 (17.0) | 1.4 (0.8—2.6) | 0.28 | 0.4 (0.2—0.9) | 0.04* |
| ≥45 | 30 (17.3) | 1.4 (0.8—2.6) | 0.26 | 0.4 (0.2—1.0) | 0.04* |
| **Income quartiles** | | | | | |
| <5,000 | 35 (35.7) | Ref | | Ref | |
| 5000—9,999 | 40 (17.9) | 0.7 (0.4—1.2) | 0.18 | 0.8 (0.4—1.4) | 0.38 |
| 10,000—14,999 | 24 (12.9) | 0.7 (0.4—1.3) | 0.26 | 1.3 (0.6—2.6) | 0.49 |
| >=15,000 | 17 (11.9) | 0.4 (0.2—0.8) | 0.01* | 0.7 (0.3—1.6) | 0.44 |
| **Level of Education** | | | | | |
| No formal education | 33 (28.4) | Ref | | Ref | |
| 6 or fewer years | 29 (19.6) | 0.9 (0.5—1.5) | 0.61 | 1.0 (0.5—1.9) | 0.96 |
| 7 to 12 years | 25 (13.9) | 0.6 (0.3—1.0) | 0.06 | 0.7 (0.3—1.3) | 0.20 |
| Higher secondary completed or college | 29 (14.1) | 0.4 (0.2—0.8) | 0.004* | 0.5 (0.2—1.0) | 0.06 |
| **Occupation** | | | | | |
| Self employed | 18 (13.1) | Ref | | Ref | |
| Employed in government or private sector | 19 (14.6) | 1.1 (0.6—2.3) | 0.73 | 1.6 (0.8—3.4) | 0.21 |
| Laborer on daily wages | 23 (27.4) | 2.5 (1.3—5.0) | 0.01* | 2.9 (1.4—6.1) | 0.006* |
| Unemployed | 33 (24.8) | 2.2 (1.2—4.1) | 0.02* | 1.8 (0.9—3.8) | 0.10 |
| Housewife or student | 23 (13.9) | 1.1 (0.5—2.1) | 0.86 | 1.5 (0.6—3.5) | 0.37 |
| **Marital Status** | | | | | |
| Never married | 25 (14.8) | Ref | | Ref | |
| Married or unmarried but living with partner | 73 (17.8) | 1.2 (0.8—2.0) | 0.39 | 1.4 (0.8—2.6) | 0.23 |
| Divorced, separated, or widowed | 18 (25.7) | 2.0 (1.0—4.0) | 0.048* | 1.6 (0.7—3.7) | 0.30 |
| **CLINICAL FACTORS** | | | | | |
| **Phase of Therapy** | | | | | |
| Intensive phase | 39 (18.0) | Ref | | Ref | |
| Continuation phase | 77 (17.8) | 1.0 (0.6—1.5) | 0.95 | 1.2 (0.8—1.9) | 0.42 |
| **Category of TB** | | | | | |
| New | 87 (17.3) | Ref | | Ref | |
| Previously treated | 29 (19.9) | 1.2 (0.7—1.9) | 0.47 | 1.3 (0.8—2.2) | 0.33 |

| | | | | | |
|---|---|---|---|---|---|
| **Type of TB** | | | | | |
| Extrapulmonary | 32 (15.8) | Ref | | Ref | |
| Smear-negative pulmonary | 67 (87.0) | 1.2 (0.7—1.9) | 0.50 | 1.4 (0.7—2.9) | 0.36 |
| Smear-positive pulmonary | 17 (4.6) | 1.5 (0.8—2.9) | 0.22 | 1.1 (0.7—1.9) | 0.66 |
| **HIV co-infection** | | | | | |
| No | 43 (12.4) | Ref | | Ref | |
| Yes | 73 (24.1) | 2.2 (1.5—3.4) | <0.001* | 2.0 (0.9—4.5) | 0.10 |
| **STRUCTURAL FACTORS** | | | | | |
| **Mode of transport to treatment site** | | | | | |
| Walking or bicycle | 26 (11.1) | Ref | | | |
| Motorcycle or car | 6 (13.6) | 1.3 (0.5—3.3) | 0.63 | | |
| Autorickshaw or taxi | 22 (16.8) | 1.6 (0.9—3.0) | 0.13 | | |
| Public transportation | 62 (25.7) | 2.8 (1.7—4.6) | <0.001* | | |
| **Money spent to collect medications** | | | | | |
| 0—24 | 30 (12.9) | Ref | | | |
| 25—49 | 15 (13.5) | 1.1 (0.5—2.1) | 0.87 | | |
| 50—75 | 24 (21.6) | 1.9 (1.0—3.4) | 0.04* | | |
| >75 | 47 (24.1) | 2.1 (1.3—3.6) | 0.003* | | |
| **Time spent to collect medication** | | | | | |
| <30 minutes | 4 (3.6) | Ref | | Ref | |
| 30 to 59 minutes | 32 (16.8) | 5.3 (1.8—15.5) | 0.002* | 5.7 (1.9—17.1) | 0.002* |
| >=60 minutes | 80 (22.9) | 7.9 (2.8—22.1) | <0.001* | 6.4 (1.9—21.4) | 0.003* |
| **PSYCHOSOCIAL FACTORS** | | | | | |
| **Current tobacco use** | | | | | |
| No | 93 (17.2) | Ref | | | |
| Smokeless tobacco use only | 7 (13.7) | 0.8 (0.3—1.8) | 0.53 | | |
| Cigarettes or beedi use | 16 (27.1) | 1.8 (1.0—3.3) | 0.07 | | |
| **Probable alcohol use** | | | | | |
| No alcohol use | 96 (16.2) | Ref | | Ref | |
| Any alcohol use | 20 (33.9) | 2.6 (1.5—4.7) | 0.001* | 2.1 (1.1—4.1) | 0.03* |

INR=Indian rupees.
[a]Proportion represents the number of participants with suboptimal adherence in a given category; for example, 49/271 females were suboptimally-adherent.

*Patient-reported reasons for non-adherence: details on reasons patients ran out of pills and medication adverse effects experienced*

Among 35 patients who reported non-adherence because they ran out of pills, 12 (34.3%) reported that this was related to difficulties in picking up medication refills (e.g., transportation challenges, illness), 8 (22.9%) forgot to pick up their medication refills, 6 (17.1%) went to pick up a refill but found that the healthcare provider was absent or the clinic was out of stock of medications, 5 (14.3%) went to pick up a refill but found the clinic was closed due to a holiday, 2 (5.7%) could not pick up medications due to travel out of town, and 2 (5.7%) reported other personal barriers to picking up medication refills. Among 29 patients who reported non-adherence due to fear of medication side effects, 10 (34.5%) had experienced nausea, 5 (17.2%) had experienced fatigue, 3 (10.3%) had experienced rash, 7 (24.1%) experienced other side effects (e.g., jaundice, giddiness, fever), and 4 (13.8%) reported a general concern that TB medications are "harmful."

*Association between non-adherence to medications and treatment card-recorded TB treatment outcomes*

Using the treatment outcomes collected on treatment cards, 381 (58.6%) patients in the cohort had treatment outcomes available, while 269 (41.4%) patients were still on treatment at the time data collection ended for the study. Patients who were lost to follow-up had statistically significantly higher odds of being non-adherent (Table S3). The outcome of death was also associated with increased odds of non-adherence, although this did not achieve statistical significance.

Table S3. Association between treatment outcomes recorded on treatment cards and medication non-adherence (N=650)

| **Treatment outcomes** | **Descriptive statistics** | | **Univariable findings** | |
|---|---|---|---|---|
| | **Proportion of overall sample in given category[a]** **N (%)** | **Proportion of medication non-adherence[b] (negative urine INH test result)** **n (%)** | **Odds ratio (95% confidence interval)** | **p-value** |
| Treatment success (cure or treatment completed) | 332 (51.1%) | 35 (10.5%) | Ref | |
| On treatment | 269 (41.4%) | 27 (10.0%) | 0.95 (0.56-1.61) | 0.84 |
| Lost to follow-up | 20 (3.1%) | 8 (40.0%) | 5.66 (2.16-14.79) | <.001* |
| Died | 15 (2.3%) | 4 (26.7%) | 3.09 (0.93-10.21) | 0.065 |
| Treatment regimen changes or transfer of care out of district | 3 (3.9%) | 3 (100.0%) | 2.31 (0.62-8.70) | 0.214 |

[a]Proportion represents the number of participants in a category divided by the overall cohort sample; for example, 332/650 participants experienced treatment success.
[b]Proportion represents the number of participants with non-adherence in a given category; for example, 35/332 participants with treatment success were non-adherent.
